# An automated EEG algorithm detect polymorphic delta activity in acute encephalopathy presenting as postoperative delirium

**DOI:** 10.1101/2022.09.01.22279475

**Authors:** Fienke L Ditzel, Suzanne CA Hut, Sandra MA Dijkstra-Kersten, Tianne Numan, Frans SS Leijten, Mark van den Boogaard, Arjen JC Slooter

## Abstract

**Aim:** Delirium, a clinical manifestation of acute encephalopathy, is often unrecognised. An important electroencephalography (EEG) characteristic of acute encephalopathy is polymorphic delta activity (PDA), which can be detected automatically. We aimed to study whether automated assessment of PDA in unselected EEG could detect acute encephalopathy that presents clinically as delirium.

**Methods:** We assessed PDA in 145 elderly patients using the first 96 seconds of unselected single-channel EEG (Fp2-Pz). We compared fully automated PDA detection with visual inspection by EEG experts. Additionally, we tested its performance as a delirium monitor by comparing PDA detection with a standardized delirium assessment by a clinical expert panel.

**Results:** PDA detection showed an area under the receiver operating characteristic (AUC) of 0.86 (95%CI 0.81–0.90) compared to EEG experts. When compared with the delirium classification of clinical experts, PDA detection achieved an AUC of 0.78 (95%CI 0.71–0.85). PDA detection correlated with the likelihood of delirium, its severity and the levels of attention and consciousness (all p<0.001).

**Conclusion:** Automated PDA detection in unselected, single-channel EEG can classify acute encephalopathy clinically presenting as delirium.

**Significance:** A fully automated EEG algorithm can assist in the recognition of delirium.

## 1. Introduction

Delirium is associated with higher mortality^1^ long-term cognitive impairment,^1–3^ longer duration of hospital admission,^4^ decline in independently living^5^ and therefore increased health care costs.^6,7^ It is highly prevalent, with an overall frequency of 23% in hospitalised patients.^8^ Recognition of delirium is essential for early treatment of underlying conditions and optimal communication, as a delirious state may impair a patient’s ability to understand explanations of procedures and prognosis.^9^

Unfortunately, delirium often remains undetected in various hospital settings.^9–12^ Therefore, several delirium assessment tools have been developed to improve recognition.^11,13^ Although excellent results have been reported in research settings, the diagnostic performance of these tools was found to be disappointing in routine, daily practice^14,15^ as key elements of these tools are subjective and therefore difficult to standardise if numerous nurses use the tool as part of daily care. Further, the diagnosis of delirium may be difficult, as experts were found to disagree in 21% of cases, even though they based their conclusion on exactly the same information.^16^ Therefore, there is a need for an objective monitor to routinely determine the brain state of patients at risk for delirium.

Acute encephalopathy is a rapidly developing pathobiological process in the brain that can be objectively measured with electroencephalography (EEG), and that clinically presents as (subsyndromal)delirium or, in severe cases, as coma.^17^ The association between delirium and EEG slowing has been known for decades.^18–20^ Since EEG slowing correlates with delirium severity,^21–23^ and normalises after delirium resolution,^24^ EEG may have the potential to monitor delirium over time. However, conventional EEG is time-consuming, expensive and unpractical for routine delirium assessments, as it can only be performed and interpreted by trained personnel.

We previously showed that single-channel EEG (Fp2-Pz) can detect postoperative delirium based on increased relative delta power,^25^ which we validated in a multicentre study.^22^ However, relative delta power can be nonspecific since noise, such as eye- and glosso-kinetic movement artefacts, also manifest predominantly within this range. Another important limitation of previous studies was the manual selection of EEG epochs at first.^18,20^ Selection of artefact-free epochs can be time consuming and has a high inter-operator variability.^26^ Therefore, results of these previous studies cannot be used in a routine clinical setting. To provide a delirium monitoring tool suitable for routine daily care, an algorithm is needed that functions without any manual EEG epoch selection after recording.

Acute encephalopathy presenting as delirium is characterised in the EEG by arrhythmic slow waves with suppression of alpha activity, known as polymorphic delta activity (PDA).^13,27–29^ Based on defined wave shape characteristics for PDA and automated detection and rejection, an algorithm was developed to function without any pre-selection of EEG epochs for delirium monitoring in daily clinical practice. The aim of this study was to test the hypothesis that a fully automated PDA wave shape algorithm can detect acute encephalopathy in unselected single-channel EEG, clinically presenting as postoperative delirium.

## 2. Methods

### 2.1 Study design and population

The current investigation is based on data from a prior prospective, multicentre cohort study, described in more detail elsewhere.^22^ The study design was approved prior to patient enrolment by the local ethical committee of University Medical Center Utrecht (protocol 13-634) and registered at clinical trial (NCT02404181, principal investigator: AJC Slooter). This manuscript adheres to the applicable Standards for Reporting of Diagnostic Accuracy Studies (STARD) guidelines. In short, elderly patients were included who were ≥ 60 years, scheduled for major surgery with an expected hospital stay of ≥ 2 days, and considered at risk of delirium.^6^ All patients gave signed informed consent and anonymity was preserved. Exclusion criteria were neurosurgery and the inability to undergo cognitive testing due to deafness or a language barrier. Additionally, we excluded patients with a Mini-Mental State Examination (MMSE) ≤ 23 because dementia could affect slow-wave EEG activity.^30,31^

### 2.2 Measurements

Patients underwent EEG recordings in resting state with eyes closed. All measurements were performed by a trained researcher on the day before surgery (T-1) and during each of the first three postoperative days (T1-T3). The researcher constantly ensured patients were awake and sat still while recording. EEG measurements were directly followed by an extensive delirium assessment, described below.

#### 2.2.1 Assessment of acute encephalopathy by EEG experts

The reference for acute encephalopathy assessment consisted of the classification of three EEG experts from the Department of Clinical Neurophysiology at the University Medical Center Utrecht, the Netherlands who, independently of each other, visually inspected the single-channel EEGs. All experts had over 15 years of clinical EEG experience. To provide training in recognising delta waves and artefacts such as eye movement, a training dataset was used,^25^ which contained of 21-channel EEGs (n=56) paired with their Fp2-Pz EEG derivation. PDA was defined when four criteria were met: 1) a frequency within 0.5 to 5 Hz; 2) being present at least three times per minute; 3) containing at least two sequent waves; 4) having a higher amplitude than alpha activity in the same recording. Blinded to clinical information, the EEG experts classified the single-channel EEGs into either “acute encephalopathy”, “possible acute encephalopathy/doubt”, or “no acute encephalopathy”. To provide a final binary conclusion “no acute encephalopathy” or “acute encephalopathy”, discussion sessions were organised to evaluate the EEGs for which there was no majority vote or when the majority vote conclusion was “doubt”.

#### 2.2.3 Assessment of delirium by clinical experts

Delirium assessments were performed by trained researcher based on a standardised, videotaped cognitive assessment of about fifteen minutes that included the Delirium Rating Scale Revised-98 (DRS-R-98),^32,33^ the Richmond Agitation and Sedation Scale (RASS),^34^ and the Confusion Assessment Method in the ICU (CAM-ICU).^34,35^ These videotapes were rated by varying pairs of two, or in case of disagreement three, delirium experts. The complete panel contained 38 clinicians, mainly psychiatrist and geriatricians, with at least five, but mostly over 10 years of experience. Blinded to each other and the EEG recording, they classified patients as either having “no delirium”, “possible delirium/subsyndromal delirium” or “delirium”, according to the Diagnostic and Statistical Manual of Mental Disorders (DSM-5) delirium criteria. In addition, the clinical experts reported the likelihood of delirium on a numeric rating scale (NRS) ranging from 1 to 10 (higher scores represented a higher likelihood of delirium). The final classification was based on the majority vote, where “possible delirium” was grouped with “delirium.”

#### 2.2.3 Detection of polymorphic delta activity

Detection of PDA was done with a fully automated wave shape analysis algorithm (DeltaScan algorithm version 2.4.2). Of the recorded channels Fp2-Pz and T8-Pz (using Fpz as a reference), analyses were solely performed on Fp2-Pz with a sampling frequency of 512Hz. The algorithm consists of 1) a pre-processing module, 2) an artefact module and 3) a module to detect PDA wave shapes.

The first pre-processing step was to use a high-pass filter (cut-off 0.125Hz) to remove low-frequency noise and slow drift from the EEG recordings. Secondly, multiple filters were used to remove line noise from power grids (50/60), device disturbances (24/64 Hz) and their harmonics. After pre-processing, the artefact module ran to reject non-EEG signals such as motion artefacts, disconnected electrodes or strong electrical interference.

Detection of PDA was run on the first non-rejected 96 seconds using a classifier based on supervised machine learning techniques. The classifier was trained to recognise wave characteristics of PDA on a training dataset that contained surgical patients with delirium (n=28) or without delirium (n=28).^25^ Training data of healthy volunteers (n=27) was added to provide target-free EEG to distinguish PDA from delta activity generated by eye movements. Both types of wave shapes were manually marked by a clinical technician with 10 years of experience in neurophysiology. Examples of these detected wave shapes and the artefact module are shown in figure 1.

**Figure 1.**
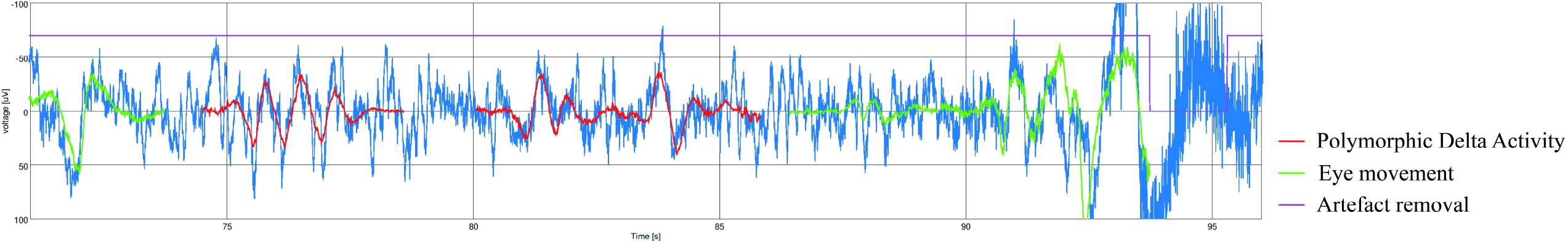
Examples of detected wave shapes. Examples of detected wave shapes including Polymorphic Delta Activity (PDA) (red) and eye movement (green). The pink line represents the artefact algorithm that deselects improper EEG signal.

Thereafter, the amount of detected PDA was translated to an ordinal score ranging from 1 to 5. This PDA Score aims to represent the likelihood of acute encephalopathy labelled: “1” very unlikely, “2” unlikely, “3” possibly, “4” likely and “5” very likely. Within this range, scores 1 and 2 are intended to represent “no acute encephalopathy” and scores 3-5 “acute encephalopathy”.

PDA Scores were set on the prespecified Receiver Operating Characteristic-curve (ROC) of our sample that was completely independent of the training dataset. A PDA Score of “1” was assigned when none or only one specific PDA wave was detected. Since the boundary between PDA Scores 2 and 3 is crucial for assignment to the categories “no acute encephalopathy” and “acute encephalopathy an optimum was chosen between the sensitivity, specificity and negative predictive value (NPV) using the EEG experts and the clinical experts described above. The boundaries between PDA Scores 3, 4 and 5, were set by sorting the positively assessed EEG measurements based on the amount of detected PDA and dividing them into three equal bins. Examples of recordings with PDA Scores (1-5) are shown in figure 2.

**Figure 2.**
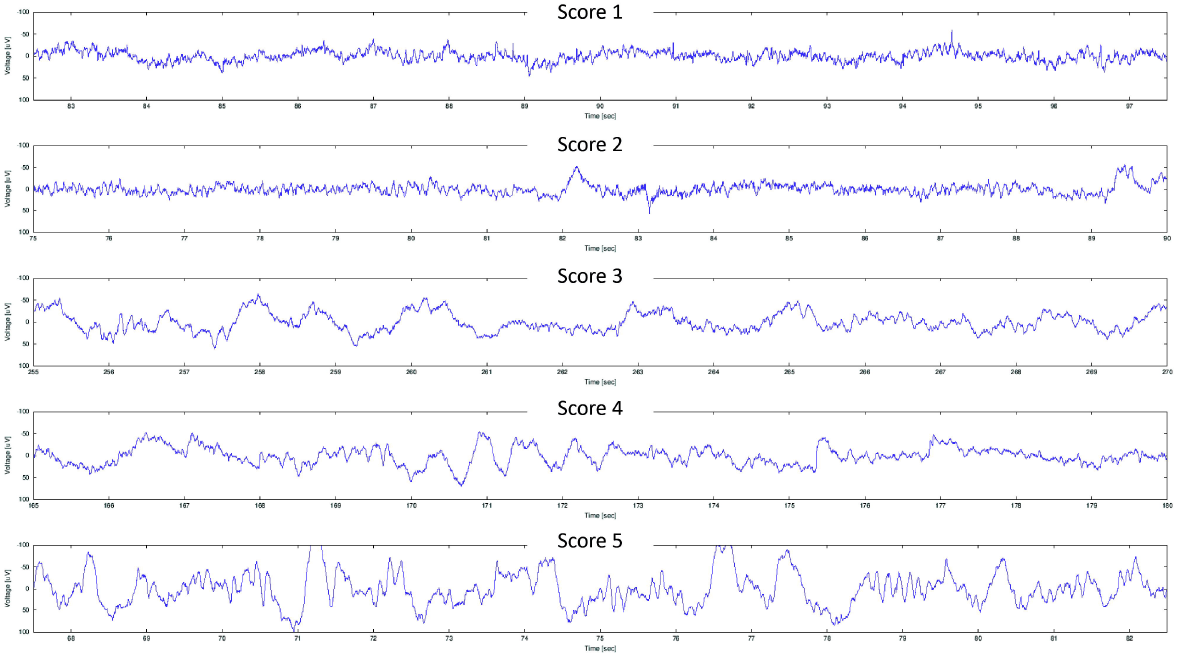
Examples of Polymorphic Delta Activity (PDA) Score. Examples of single-channel EEG recordings (Fp2-Pz) of each PDA Score ranging from 1 (no PDA) to 5 (a lot of PDA). This PDA Score aims to represent the likelihood of acute encephalopathy labelled: “1” very unlikely, “2” unlikely, “3” possibly, “4” likely and “5” very likely. Within this range, scores 1 and 2 are intended to represent “no acute encephalopathy” and scores 3-5 “acute encephalopathy”.

### 2.3 Statistical analyses

First, we compared positive assessments for acute encephalopathy as classified by PDA detection (PDA Score 3-5) with acute encephalopathy according to EEG experts. Secondly, we compared these positive assessments with assessments classified as delirium according to the clinical experts. Next, we expressed the performance of PDA detection as the Area Under the Receiver Operating Characteristic (AUC) for the two expert panels. In addition, we performed a stratified analysis based on the presence or absence of a medical history of stroke or transient ischemic attack (TIA) and compared strata with DeLong’s test.

Predictive values were calculated for each PDA Score boundary ranging from 1-5. Spearman’s rank correlation coefficients (r_s_) were determined to investigate correlations between PDA Scores (1-5) and the scores of the clinical experts on likelihood of delirium (averaged Numeric Rating Scale, NRS), the severity of delirium (averaged DRS-R-98), level of attention (i.e., averaged item-10 of the DRS-R-98 score), and level of consciousness (averaged RASS). A P-value of < 0.05 was considered significant. Analyses were performed with SPSS version 26.0.0.1 and Ri386 version 4.0.3.

## 3. Results

### 3.1 Study population

Of the 159 patients (n=360 assessments) that were available for analysis, we excluded 10 patients (n=21 assessments) with a preoperative MMSE score ≤ 23 from the analysis. Subsequently, we excluded assessments with missing expert diagnoses due to incomplete clinical data (n=7 assessments) or insufficient EEG (n=11 assessments). Of the remaining 145 patients (n=321 assessments), the automated artefact algorithm rejected 9 assessments (success rate 97%). A flowchart of the included patients and assessments can be found in Supplement 1. The final study population included 145 patients (n=312 assessments). Patient characteristics are shown in Table 1.

**Table 1.** Patient characteristics.

|  | **All patients (n=145)** |
| --- | --- |
| **Age in years, mean (SD)** | 77 (6.3) |
| **Male sex, n (%)** | 99 (68%) |
| **Female sex, n (%)** | 46 (32%) |
| **MMSE<sup>†</sup>, median (IQR)** | 28 (27-29) |
| **Medical history, n (%)** |  |
| **Stroke or TIA<sup>‡</sup>** | 40 (29%) |
| **Any psychiatric disease<sup>§</sup>** | 6 (4%) |
| **>10 IE alcohol/week** | 29 (25%) |
| **Alcohol IE/week, median (IQR)** | 2 (12) |
| **Benzodiazepine use <24h** | 25 (17%) |
| **Surgery type, n (%)** |  |
| **Cardiothoracic or vascular** | 131 (90%) |
| **Orthopaedic** | 8 (6%) |
| **Other** | 6 (4%) |

*Data are presented as mean with standard deviation (SD), median with interquartile range (IQR), or number (n) in percentage (%).<sup>†</sup>Mini-Mental State Exam, <sup>‡</sup>Transient ischemic attack, <sup>§</sup>All patients with a psychiatric disease had a medical history of depression. One patient had a medical history of bipolar disorder.*

### 3.2 Classification of delirium/acute encephalopathy

The EEG experts classified 47% of the population with acute encephalopathy (68 patients, 115 assessments) and the clinical experts diagnosed 32% of the patients with delirium (47 patients, 68 assessments). The overall overlap between both expert panels on assessment level was 70%. No patients were classified as delirious on the day before surgery (T-1) by the clinical experts. PDA detection distinguished five groups with the following labels: “1” very unlikely (n=172 assessments, 55%), “2” unlikely (n=25 assessments, 8%), “3” possibly (n=47 assessments, 15%), “4” likely (n=37 assessments, 12%) and “5” very likely (n=31 assessments, 10%). An overview of these groups and classification of both panels can be found in Supplement 2.

### 3.3 Accuracy of polymorphic delta detection

The AUC for PDA detection was 0.86 (95% CI 0.81–0.90) for acute encephalopathy according to the EEG experts, and 0.78 (95% CI 0.71– 0.85) for delirium according to the clinical experts (Figure 3).

**Figure 3.**
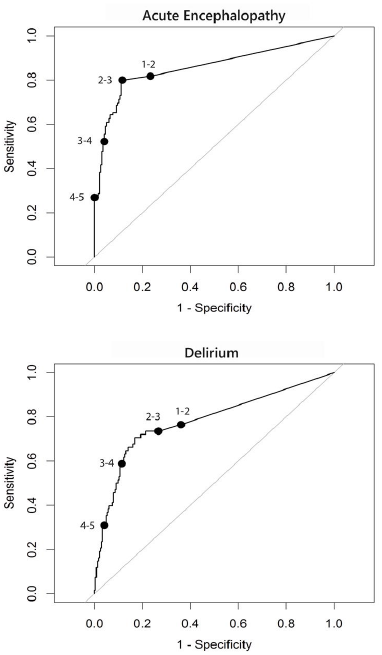
Receiver Operating Characteristic (ROC) curves for Polymorphic Delta Activity (PDA) ROC-curves show the sensitivity and specificity for every boundary of the PDA Score. The Area Under the Receiver Operating Characteristic (AUC) for acute encephalopathy was 0.86 (95% Confidence Interval (CI) 0.81-0.90) using visual inspection of EEG experts as a reference. The AUC for delirium was 0.78 (95% CI 0.71-0.85) using clinical experts as a reference.

The boundary between 2 and 3 on the ROC, representing the cut off between acute encephalopathy and no acute encephalopathy, showed a sensitivity of 0.80 and a specificity of 0.88, according to the EEG experts. The same boundary showed a sensitivity of 0.74 and a specificity of 0.73 for delirium according to the clinical experts. Table 2 shows the predictive values for all PDA Scores. A specificity of 1 was reached for a PDA Score of “5” compared to visual inspection of EEG experts and 0.96 for the clinical reference.

**Table 2.**
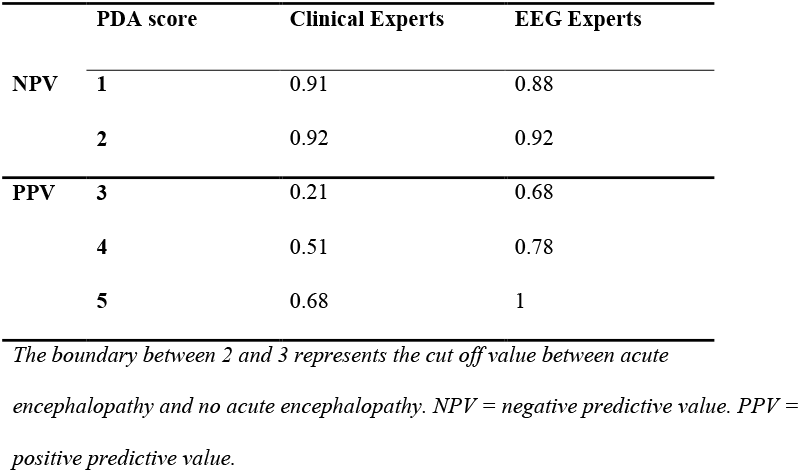
Predictive values for each Polymorphic delta activity (PDA) Score.

|  | **PDA score** | **Clinical Experts** | **EEG Experts** |
| --- | --- | --- | --- |
| **NPV** | **1** | 0.91 | 0.88 |
|  | **2** | 0.92 | 0.92 |
| **PPV** | **3** | 0.21 | 0.68 |
|  | **4** | 0.51 | 0.78 |
|  | **5** | 0.68 | 1 |

*The boundary between 2 and 3 represents the cut off value between acute encephalopathy and no acute encephalopathy. NPV = negative predictive value. PPV = positive predictive value.*

When we stratified our data based on the presence or absence of a previous stroke or TIA, there was no significant difference in the AUC using either expert panel (Supplement 3).

The PDA Score (1-5) correlated significantly with the likelihood of delirium (NRS, r_s_=.37, 95%CI 0.25-0.47), the severity of delirium (DRS-R-98, r_s_ = .46, 95%CI 0.36-0.55), the level of attention (i.e. item-10 of the DRS-R-98 score, r_s_ = .41, 95%CI 0.32-0.51) and level of consciousness (RASS, r_s_= -.32, 95%CI -0.44- -0.19) (p < .001 for all comparisons).

## 4. Discussion

This multicentre cohort study aimed to investigate the diagnostic performance of PDA detection on manually unselected single-channel EEG in diagnosing acute encephalopathy presenting as postoperative delirium. In summary, PDA detection classified acute encephalopathy with an AUC of 0.86 compared to visual inspection by EEG experts. An AUC of 0.78 was found for delirium detection using the clinical experts as a reference. PDA detection correlated with the likelihood and severity of delirium as well as the levels of attention and consciousness. Importantly, this performance is similar to automated detection of relative delta power in single-channel EEG that was manually selected for artefact-free epochs,^22^ hampering application in routine, daily care.

Interestingly, we found that the EEG experts included more positive assessments than the clinical experts (115 and 68, respectively). EEG abnormalities accompanying delirium have shown to been associated with poor outcomes, such as mortality and length of stay.^36,37^ Previously, we showed that acute encephalopathy in EEG without confirmed delirium is associated with a significantly higher DRS-R-98 score than no acute encephalopathy and no delirium.^29^ In the current study, the amount of PDA correlated with delirium severity. We speculate that assessments classified as acute encephalopathy without delirium, indicate subsyndromal delirium. Such subsyndromal delirium could resolve before it becomes clinically apparent or could deteriorate to full-blown delirium. PDA may therefore be an early indicator of delirium, allowing earlier treatment of underlying conditions.^38,39^

Interest in minimal lead EEG-based delirium monitoring seems to be rising. Previous studies showed a similar performance to the findings in this study^37,40,41^ but did not use a pre-hand trained automated artefact algorithm. For application in clinical practice, automated artefact detection and rejection is an important feature. Secondly, previous studies had a data-driven approach instead of validating findings^25^ in a completely independent dataset as was done in this study.

An important strength of our study was furthermore the use of a comprehensive reference standard, including two expert panels. PDA detection was compared with the classification of three experienced, well-trained EEG experts assessing all EEG. In addition, PDA detection was compared with the classification of two (or three, in case of discordance) experienced clinicians, who based their diagnosis on at least 20 minutes of video recorded cognitive testing. It is essential that reference panels consist of more than one expert, as we showed previously that experts often disagree on the diagnosis of delirium although they based their conclusion on exactly the same clinical information.^16^ Since the cognitive assessment and EEG measurement were immediately followed by each other, we were able to capture the patient’s status both clinically and physiologically in a time frame that is unlikely to be subject to fluctuations.

A limitation of the study is the inclusion of only postoperative patients, mainly after cardiothoracic or vascular surgery. The inclusion of a more heterogeneous group of patients would allow us to determine whether the results can be generalised to other settings. Our proper screening of patients and videotaped cognitive testing make it unlikely - although not impossible - that any present PDA could be due to other causes. For example, an unknown structural cerebral abnormality^42^ could possibly affect the outcomes. However, analyses that were stratified on the presence or absence of a previous stroke or TIA, yielded similar results. Delta activity due to sleep seems unlikely since the researcher constantly ensured that patients were awake.

Further research should explore the properties of the studied algorithm in patients who may show increased delta activity in EEG, such as dementia,^30,31,43^ or other neurological^42^ and psychiatric disorders.^44,45^ Secondly, since the presence and severity of delirium symptoms may change within 24 hours, it is important to study the fluctuation of delirium symptoms over the day and their correspondence with detection of PDA. Lastly, the correlation between PDA and length of stay or mortality is important to understand the significance of EEG based monitoring. PDA detection can be implemented in daily clinical care, since output is generated without any manual pre-selection of epochs after recording. The technology presented can be incorporated into an easy-to-apply tool with a simple likelihood score (1-5) to measure altered mental status. PDA detection can especially aid in the diagnosis of hypoactive delirium, which is more common but more often no recognised than hyperactive delirium and may be confused with fatigue, sleepiness or depression.^46^ The PDA score allows for the monitoring of brain function as a time dependent parameter such as temperature and blood pressure.

In conclusion, since delirium is often not detected in usual care, recognition could be improved with an easy-to-apply EEG monitor with automated analysis. Our findings show that automated detection of PDA can diagnose acute encephalopathy clinically presenting as delirium.

## Data Availability

All data produced in the present study are available upon reasonable request to the authors

## Highlights

- We validated a fully automated algorithm with an integrated artefact detector based on prespecified wave shape characteristics
- Detection of polymorphic delta activity was compared with visual inspection of EEG and delirium assessment based on 20-minute videotaped cognitive testing
- The amount of PDA can be translated in a delirium probability score that can be used in daily delirium monitoring with single-channel EEG

## Acknowledgements

The authors would like to especially thank The Dutch delirium study group who served as members of the clinical expert panel, as well as Nizare R V R Henriquez and Nico W Teunissen of the Department of Neurology and Neurosurgery and University Medical Center Utrecht Brain Center, University Medical Center Utrecht, Utrecht University, Utrecht, The Netherlands, who served as members of EEG expert panel.

## Disclosure

The sponsor and Prolira, the company that developed the monitor and the latest algorithm in detecting PDA, had no role in the study design, data analysis, data interpretation, writing of the report, or the decision to submit for publication.

## Declaration of interests

Arjen JC Slooter is an advisor for Prolira, a start-up company that develops an EEG-based delirium monitor. Any (future) profits from EEG-based delirium monitoring will be used for future scientific research only. None of the other authors reports any conflicts of interest.

## Funding

This work was supported by European Union Horizon 2020 [grant number 820555].

## Details of authors’ contribution

All authors confirmed they have contributed to the intellectual content of this article and gave final approval of the version to be published.

<u>Fienke L Ditzel</u>: this author contributed with data preparation, analysis and preparation of the manuscript

<u>Suzanne CA Hut</u>: this author contributed with data preparation, analysis and revision of the manuscript

<u>Sandra MA Dijkstra-Kersten</u>: this author contributed with data preparation and analysis

<u>Tianne Numan:</u> this author contributed with data collection, data preparation, revision of the manuscript

<u>Frans SS Leijten:</u> this author contributed with data collection and revision of the manuscript

<u>Mark van den Boogaard</u>: this author contributed with data collection and revision of the manuscript

<u>Arjen JC Slooter:</u> this author contributed with data collection and revision of the manuscript

## Glossary of Terms

AUC: Area Under the receiver operating Characteristic
BMI: Body Mass Index
CAM-ICU: Confusion Assessment Method in the ICU
CAM-S: Confusion Assessment Method-Severity
CI: Confidence Interval
DSM-5: Diagnostic and Statistical Manual of Mental Disorders fifth
DRS-R-98: Delirium Rating Scale Revised-98
EEG: Electroencephalogram
IQR: Interquartile range
MMSE: Mini-Mental State Examination
NPV: Negative Predictive Value
NRS: Numeric Rating Scale
PPV: Positive Predictive Value
RASS: Richmond Agitation and Sedation Scale
ROC: Receiver Operating Characteristic-curve
SD: Standard Deviation
TIA: Transient Ischemic Attack

## Supporting information

1. Flowchart of included patients and assessments
2. Polymorphic delta activity (PDA) Score compared to the expert panel
3. Stratified analysis for a medical history containing TIA or Stroke

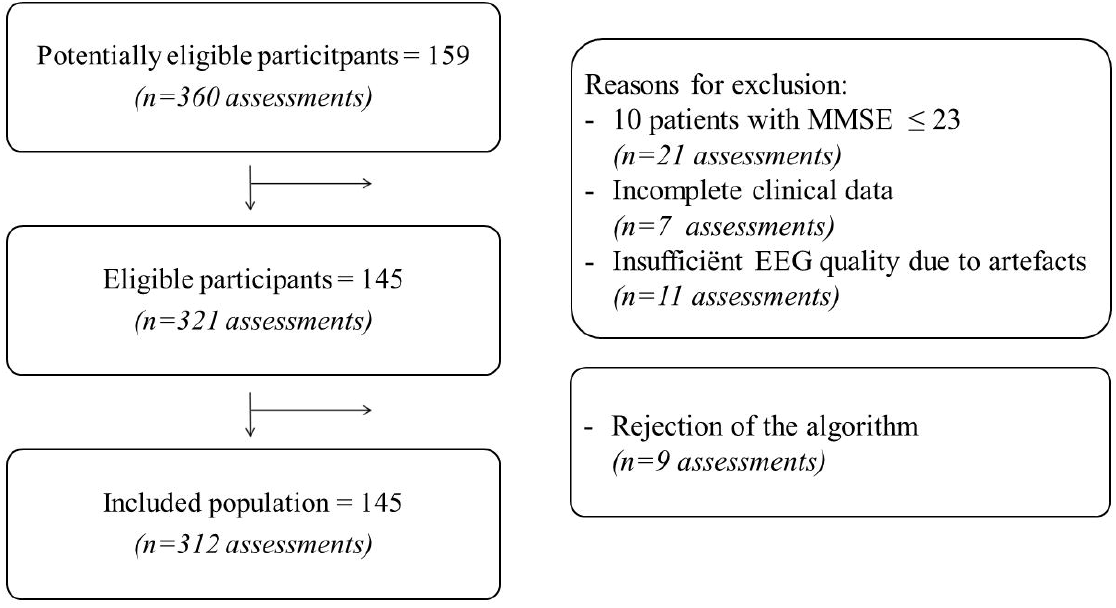

## Notes

### Competing Interest Statement

Arjen JC Slooter is a non-salaried advisor for Prolira, a start-up company that develops an EEG-based delirium monitor. Any (future) profits from EEG-based delirium monitoring will be used for future scientific research only. Frans SS Leijten is also a non-salaried advisor and holds shares in Prolira. None of the other authors reports any conflicts of interest. The sponsor and Prolira, had no role in the study design, data analysis, data interpretation, or the decision to submit for publication.

### Clinical Trial

NCT02404181

### Funding Statement

This work was funded by European Union Horizon 2020 [grant number 820555].

### Author Declarations

The study design was approved prior to patient enrolment by the local ethical committee of University Medical Center Utrecht (protocol 13-634)

