## Supporting information for "An automated EEG algorithm detect polymorphic delta activity in acute encephalopathy presenting as postoperative delirium"

### Supplement 1 Flowchart of included patients and assessments

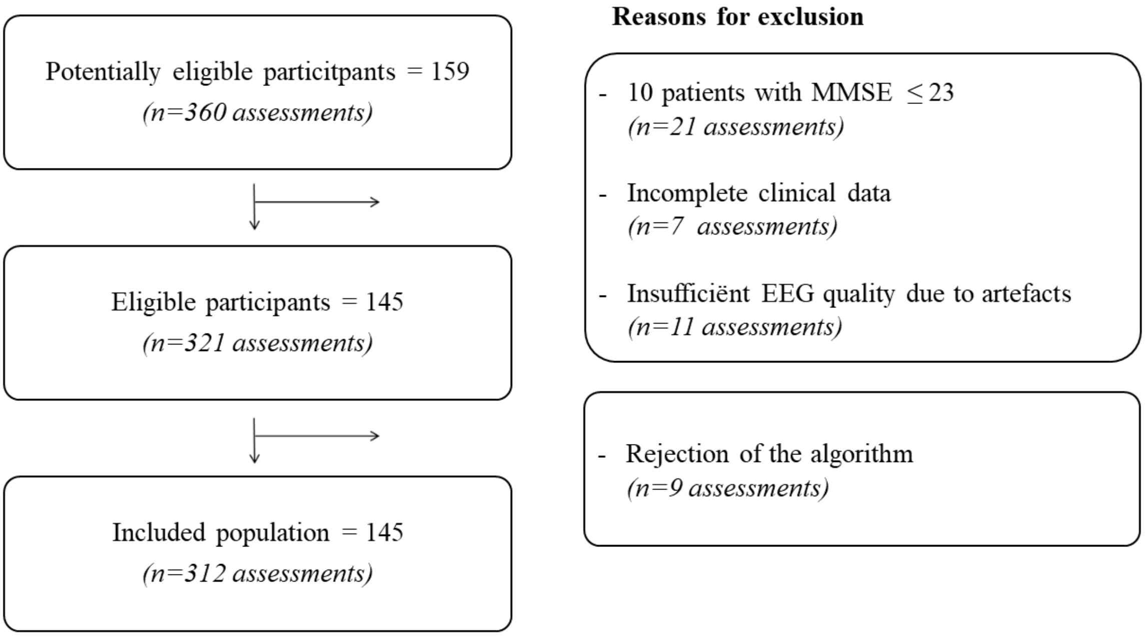


*Patients with Mini Mental State examination (MMSE)<23 were excluded, for dementia could affect slow-wave EEG activity. Assessments with insufficient EEG data due to artefacts were selected by the EEG reference.*

### Supplement 2 Polymorphic delta activity (PDA) Score compared to the expert panels

| PDA | DRS-98-R | RASS | Acute encephalopathy | | Delirium | | | Total |
|---|---|---|---|---|---|---|---|---|
| Score | Median (IQR) | Median (IQR) | **-** | + | **-** | **+** (hypo/mixed/hyper) | | |
| **1** | 2.5 (1.1 - 3.9) | 0 (0 - 0) | 151 | 21 | 156 | 16 | (6 / 4 / 6) | **172** |
| **2** | 3.0 (1.7 - 4.3) | 0 (0 - 0) | 23 | 2 | 23 | 2 | (0 / 1 / 1) | **25** |
| **3** | 3.5 (1.4 - 5.6) | 0 (0 - 0) | 15 | 32 | 37 | 10 | (1 / 0 / 9) | **47** |
| **4** | 6.3 (3.0 - 9.7) | 0 (-0.5 - 0.5) | 8 | 29 | 18 | 19 | (2 / 4 / 13) | **37** |
| **5** | 9.5 (4.8 - 14.2) | -1 (-1.5 - 0.5) | 0 | 31 | 10 | 21 | (11 / 1 / 9) | **31** |
| **Total** | **3.5 (1.5 - 5.5)** | **0 (0 - 0)** | **197** | **115** | **244** | **68** | **(20 / 10 / 38)** | **312** |

*Acute encephalopathy -/+ refers to the classification of the panel of EEG experts.*

*Delirium -/+ refers to the diagnosis of the panel of clinical experts.*

### Supplement 3 Stratified analysis for a medical history containing TIA or Stroke

| | AUC Acute Encephalopathy | AUC Delirium |
|---|---|---|
| Patients with a medical history containing TIA or stroke (N=71) | 0.81 (0.71-0.91) | 0.82 (0.71-0.93) |
| Patients without a medical history containing TIA or stroke (N=213) | 0.90 (0.85-0.91) | 0.76 (0.68-0.85) |
| P-value | 0.13 | 0.42 |
